# Association of County-Wide Mask Ordinances with Reductions in Daily CoVID-19 Incident Case Growth in a Midwestern Region Over 12 Weeks

**DOI:** 10.1101/2020.10.28.20221705

**Authors:** Enbal Shacham, Stephen Scroggins, Matthew Ellis, Alexander Garza

## Abstract

**Importance:** This study assessed the longitudinal impact of new COVID-19 cases when a mask ordinance was implemented in 2 of a 5-county Midwestern U.S. metropolitan region over a 3-month period of time. Reduction in case growth was significant and reduced infection inequities by race and population density.

**Objective:** The objective of this study was to assess the impact that a mandatory mask wearing requirement had on the rate of COVID-19 infections by comparing counties with a mandatory policy with those neighboring counties without a mandatory masking policy.

**Design:** This was a quasi-experimental longitudinal study conducted over the period of June 12-September 25, 2020.

**Setting:** This study was a population-based study. Data were abstracted from local health department reports of COVID-19 cases.

**Participants:** Raw cases reported to the county health departments and abstracted for this study; census-level data were synthesized to address county-level population, income and race.

**Intervention(s) (for clinical trials) or Exposure(s) (for observational studies):** The essential features of this intervention was an instituted mask mandate that occurred in St. Louis City and St. Louis County over a 12 week period.

**Main Outcome(s) and Measure(s):** The primary study outcome measurement was daily COVID-19 infection growth rate. The mask mandate was hypothesized to lower daily infection growth rate.

**Results:** Over the 15-week period, the average daily percent growth of reported COVID-19 cases across all five counties was 1.81% (±1.62%). The average daily percent growth in incident COVID-19 cases was similar between M+ and M- counties in the 3 weeks prior to implementation of mandatory mask policies (0.90% [±0.68] vs. 1.27% [±1.23%], respectively, p=0.269). Crude modeling with a difference-in-difference indicator showed that after 3 weeks of mask mandate implementation, M+ counties had a daily percent COVID-19 growth rate that was 1.32 times lower, or a 32% decrease. At 12 weeks post-mask policy implementation, the average daily COVID-19 case growth among M- was 2.42% (±1.92), and was significantly higher than the average daily COVID case growth among M+ counties (1.36% (±0.96%)) (p<0.001). A significant negative association was identified among counties between percent growth of COVID-19 cases and percent racial minorities per county (p<0.001), as well as population density (p<0.001).

**Conclusions and Relevance:** These data demonstrate that county-level mask mandates were associated with significantly lower incident COVID-19 case growth over time, compared to neighboring counties that did not implement a mask mandate. The results highlight the swiftness of how a mask ordinance can impact the trajectory of infection rate growth. Another notable finding was that following implementation of mask mandates, the disparity of infection rate by race and population density was no longer significant, suggesting that regional-level policies can not only slow the spread of COVID-19, but simultaneously create more equal environment.

**Key Points:** *Question:* How are local mask ordinances associated with growth of COVID-19 cases among adjacent counties?

*Findings:* Ecological longitudinal analysis reveals a significant slowing of daily COVID-19 case growth after mask ordinance implementation among counties.

*Meaning:* Local-level policy of mask ordinances are shown to be an effective COVID-19 mitigation strategy even within locations of diverse populations.

## Introduction

COVID-19 has claimed over 210,000 lives in the U.S.^1^ While urgent policies have been implemented to reduce COVID-19 infections throughout the pandemic, there has been significant variability across state and local governments. In particular, due to the difficulty in maintaining social distancing guidelines and in an attempt to allow the economy to recover by opening businesses, some elected officials have issued mask mandates for the public to reduce COVID-19 transmission. Research has demonstrated the effectiveness of masks in preventing the spread of infectious diseases through airborne droplets,^2^ yet the majority of a community would need to be masked in order to reduce infection rates. Several governors have avoided state-level mandates by rationalizing that within-state variations of COVID-19 infection rates call for more localized public health policymaking than state-wide orders.^3^

While studies found mask policies to be effective in reducing infection growth rates early in the pandemic, there is an urgent need to know how the impact of such policy persists over time; in addition, the impact on unequal infection rates such as higher rates reported among African-American/Black and Hispanic/Latinx populations.^1^ Mask wearing has transitioned to a politically nuanced behavior, and this has been particularly true in the region surrounding the Midwestern city of St. Louis, providing a natural experiment to understand the longitudinal impact of a variable mask mandate policies has on county-level changes in incident CoVID-19 rates over a 12-week period.

## Methods

This ecological study evaluated the effects of a public mask mandate on the daily cumulative case growth of COVID-19 infections among five neighboring counties within the metropolitan statistical area of Saint Louis, Missouri: City of St. Louis, St. Louis County, Jefferson County, Saint Charles County, and Franklin County. The study period included a three-week period prior to the implementation of a mandatory mask order in St. Louis City and County on July 3, 2020, and a 12-week post-implementation period, for a total of 15 weeks.

### Outcome of Interest

The primary outcome of interest was the cumulative daily percent change of reported COVID-19 cases by county. Daily incident COVID-19 cases were sourced from publicly available data provided by the Missouri Department of Health and Senior Services.

### Primary Predictor and Covariates

Presence of a mask ordinance was the primary predictor of interest. Among the five counties, two implemented a mask ordinance during the study period for individuals entering public indoor spaces; the City and County of Saint Louis^4^ (M+), with three counties having no such mandate during the study period (M-).

As COVID-19 infections occur more often among areas with higher county-level population density per square mile, higher proportion of residents identifying as non-white, and lower median annual household income,^25,6^ these variables were included from the 2018, 5-year American Community Survey to control for variations in COVID-19 case growth.

### Statistical Analysis

Data were constructed with daily percent changes in COVID-19 cases as a function of time (daily) and location (county), with 530 unique observations from June 12, 2020 to September 25, 2020. The average percent change in cases per day was calculated across M+ and M- counties to understand the pattern of COVID-19 growth throughout the study period. Kendal’s tau-b rank correlation test was employed to identify associations among county-level demographics and percent daily growth of COVID-19 cases.

A difference-in-difference (DID) framework was employed to evaluate the effect of mask ordinances on percent daily growth in COVID-19 prevalence. Specific to this study, a DID estimator (δ) was calculated using binary dummy variables for both treatment (M+ or M-) at time (prior to mask ordinance implementation and after implementation). The calculated DID estimator was then applied to a linear probability model to determine the effects of a mask ordinance on COVID-19 case growth. Four adjusted models were constructed, accounting for county-level characteristics, each at 3 week intervals past the implementation date.

Statistical analyses were completed using R 4.0 software with significance determined at α = 0.05.

## Results

Over the 15-week period, the average daily percent growth of reported COVID-19 cases across all five counties was 1.81%(±1.62%). A total of 44,294 cases were reported throughout this period among a total estimated population of just over 2,000,000 residents, which is approximately one-third of the population within the state of Missouri.

The average daily percent growth in incident COVID-19 cases was similar between M+ and M- counties in the 3 weeks prior to implementation of mandatory mask policies (0.90%[±0.68] vs. 1.27%[±1.23%], respectively, p=0.269) (Figure 1). Prior to the mandate implementation a there was no association between percent growth of COVID-19 cases and percent racial minorities, population density, nor median income per county.

**Figure 1.**
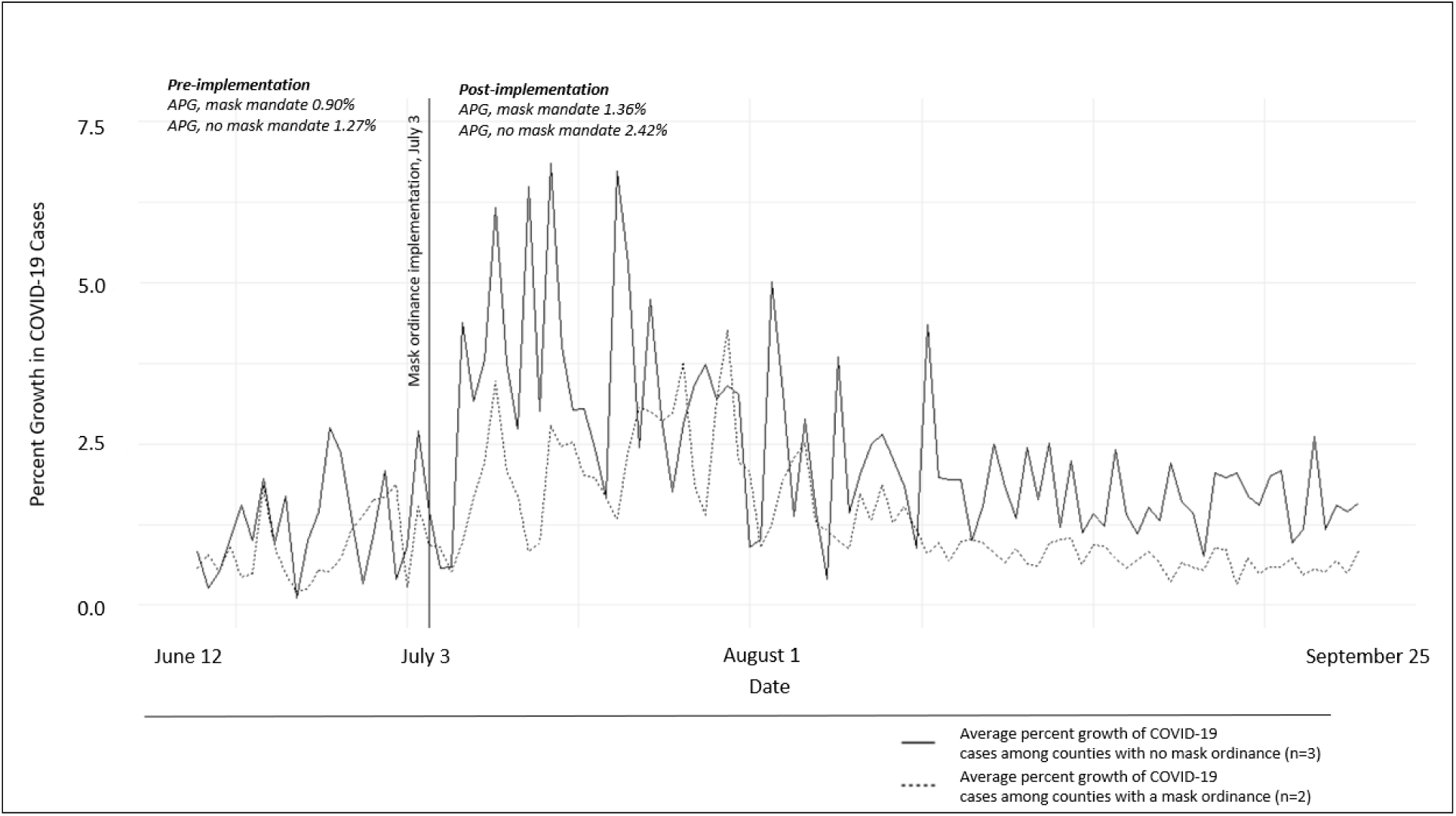
Daily average percent growth of reported COVID-19 cases among counties that implemented a mask ordinance on July 3 and those that did not.

At 12 weeks post-implementation, the average daily COVID-19 case growth among M- was 2.42%(±1.92), significantly higher than the average daily COVID case growth among M+ counties (1.36%[±0.96%], p<0.001).

In adjusted linear probability models (Table 1), the mask mandate was found to be significantly associated with slowing the reported growth of daily COVID-19 cases across the study period. From 3 weeks pre-policy to 3 weeks post-policy, M+ counties had a daily average COVID-19 growth rate of 44% less than M- counties (1.08/2.44). At week 6, 9, and 12 assessments, modeling revealed a continuing significant reduction in COVID-19 cases in counties with mask mandates. From pre-policy to 12 weeks post-policy (Figure 2), M+ counties had a daily average COVID-19 growth rate of 40% less than of M- counties (0.47/1.16). Using the DID linear probability equation, a counterfactual line is estimated, indicating probable growth of counties with mask mandate had a mask mandate not been implemented. Within the adjusted models, accounting for the DID estimator, proportion non-white population and population density were negatively associated with increased daily growth rate, yet median income did not contribute significantly to any of the calculated models.

**Table 1.** Covariate associations 3 weeks prior to mask mandate and 12 weeks after mask mandate^a^.

|  |  |  |  |  |
| --- | --- | --- | --- | --- |
|  | 3 weeks prior to mask<br>mandate, June 12 to July 3 |  | 12 weeks prior to mask mandate,<br>July 3 to September 25 |  |
| | $\tau$ | p value | $\tau$ | p value |
| Percent population non-white | -0.04 | 0.623 | -0.22 | <0.001 |
| Median annual household income (\$) | -0.01 | 0.849 | <0.01 | 0.977 |
| Population density (mi<sup>2</sup>) | -0.08 | 0.282 | -0.26 | <0.001 |
| a. Kendal's tau-b rank correlation |  |  |  |  |

**Table 2.** Linear probability models predicting percent growth in reported COVID-19 cases among five counties in Missouri.

|  | 3 Weeks Post Mask Policy |  | 6 Weeks Post Mask Policy |  | 9 Weeks Post Mask Policy |  | 12 Weeks Post Mask Policy |  |
| --- | --- | --- | --- | --- | --- | --- | --- | --- |
| | $\beta$ | Std. E | $\beta$ | Std. E | $\beta$ | Std. E | $\beta$ | Std. E |
| DID estimator<sup>a</sup> | -1.36** | 0.45 | -0.77* | 0.40 | -0.72* | 0.36 | -0.69* | 0.33 |
| Percent population non-white<sup>b</sup> | 0.10 | 0.32 | -0.08 | 0.27 | -0.07 | 0.22 | -0.04 | 0.19 |
| Median annual household income(\$)<sup>c</sup> | <0.01 | <0.01 | <0.01 | <0.01 | <0.01 | <0.01 | <0.01 | <0.01 |
| Population density (mi<sup>2</sup>)<sup>d</sup> | <0.01 | <0.01 | <0.01 | <0.01 | <0.01 | <0.01 | <0.01 | <0.01 |

a. adjusted R<sup>2</sup> 0.32, b. adjusted R<sup>2</sup> 0.21, c. adjusted R<sup>2</sup> 0.15, d. adjusted R<sup>2</sup> 0.13
Significance reported t \*0.05, \*\*0.01, \*\*\*0.001

**Figure 2.**
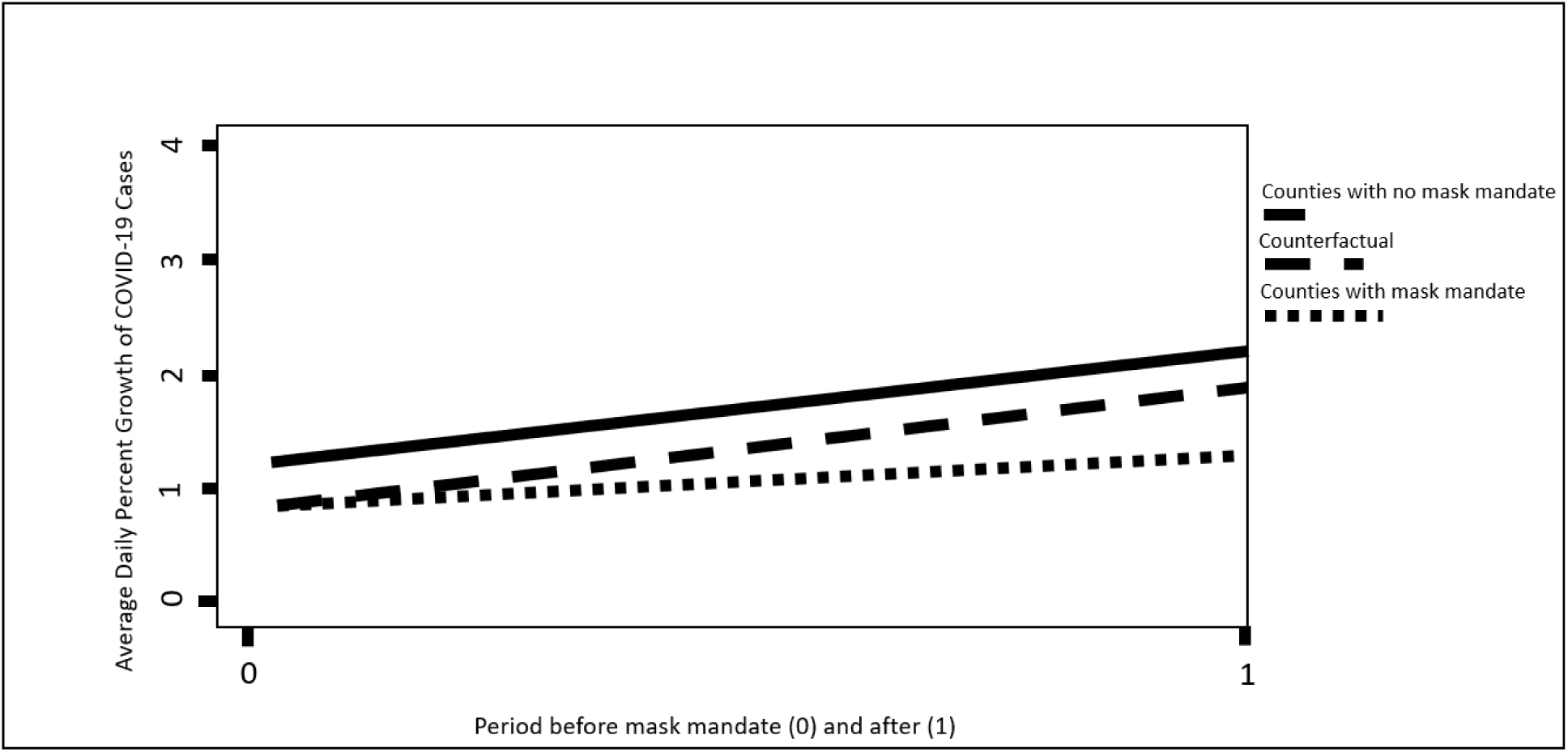
Plotting of *Y=β0+ β1[time]+ β2[presence of mask ordinance]+ β3[DID estimator]+ ε* from model results in table 1. Effect of a mask mandate among counties is depicted in the difference between the dotted mask line and the dotted/dashed counterfactual line.

## Discussion

This longitudinal study demonstrates that the implementation of county-level mask mandates lowered incident COVID-19 case growth over time, compared to neighboring counties with no such mask mandate. Importantly, the results highlight the swiftness of such impact, with significant effects seen at just 3 weeks post-implementation. As time progressed, this impact was slightly reduced, possibly due to the arbitrary political borders of counties and states, which do not necessarily constrain transmission as mobility across mask mandate and non-mandated regions occurs. Although broader state or national mandatory mask ordinances would likely have a significant reduction in COVID-19 infections, in their absence, support for governors and elected officials to minimize the interactions across these borders may be beneficial.

Furthermore, implementation of a mask mandate may have reduced the unequal burden on African American/Black and Hispanic/Latinx individuals, as well as areas with higher population density. COVID-19 has highlighted the impact and swiftness of racial and ethnic inequities through higher rates of COVID-19 morbidity and mortality among non-white communities. In particular, many individuals were unable to abide by stay-at-home orders that protected middle and high-income communities as a result of “essential” employment in grocery stores, health care support positions, and public transportation. These jobs, often lacking PPE and paid sick leave, are more often filled by people of color, living in higher population density and lower socioeconomic areas.^7^ The mask policy that was enacted in many communities may have provided a more equal approach to reducing COVID-19 infections, as it occurred in St. Louis, Missouri.

## Data Availability

Access to data is freely available from the Missouri Department of Health and Senior Services, New York Times, and U.S. Census Bureau.

